# Genome sequencing as a first-line diagnostic test for hospitalized newborns

**DOI:** 10.1101/2021.08.31.21262633

**Authors:** Kevin M. Bowling, Michelle L. Thompson, Candice R. Finnila, Susan M. Hiatt, Donald R. Latner, Michelle D. Amaral, James M.J. Lawlor, Kelly M. East, Meagan E. Cochran, Veronica Greve, Whitley V. Kelley, David E. Gray, Stephanie A. Felker, Hannah Meddaugh, Ashley Cannon, Amanda Luedecke, Kelly E. Jackson, Laura G. Hendon, Hillary M. Janani, Marla Johnston, Lee Ann Merin, Sarah L. Deans, Carly Tuura, Heather Williams, Kelly Laborde, Matthew B. Neu, Jessica Patrick-Esteve, Anna C.E. Hurst, Jegen Kandasamy, Wally Carlo, Kyle B. Brothers, Brian M. Kirmse, Renate Savich, Duane Superneau, Steven B. Spedale, Sara J. Knight, Gregory S. Barsh, Bruce R. Korf, Gregory M. Cooper

**Affiliations:** HudsonAlpha Institute for Biotechnology, Huntsville, AL; University of Alabama in Huntsville, Huntsville, AL; Children’s Hospital New Orleans, New Orleans, LA; University of Alabama at Birmingham, Birmingham, AL; University of Louisville, Louisville, KY; University of Mississippi Medical Center, Jackson, MS; Woman’s Hospital, Baton Rouge, LA; University of Utah Health, Salt Lake City, UT

**Keywords:** genome sequencing, newborns, utility, diagnostic yield, genetic diagnosis

## Abstract

**Purpose:** SouthSeq, a translational research study to perform genome sequencing (GS) for infants with symptoms suggestive of a genetic disorder, was conducted in NICUs in the Southeastern US. Recruitment targeted racial/ethnic minorities and rural, medically underserved areas that are historically under-represented in genomic medicine research.

**Methods:** GS and analysis were performed for 367 newborns to detect disease-causal genetic variation concurrent with standard of care evaluation and testing.

**Results:** Definitive diagnostic (DD) or likely diagnostic (LD) genetic findings were identified in 30% of newborns and 14% harbored an uncertain result. Only 39% of DD/LD findings were identified via concurrent standard of care suggesting that GS testing is better for obtaining early genetic diagnosis. We also identified phenotypes that correlate with the likelihood of receiving a DD/LD finding, such as craniofacial, ophthalmologic, auditory, skin, and hair abnormalities. We did not observe any differences in diagnostic rates between racial/ethnic groups.

**Conclusion:** We describe one of the largest to-date GS cohorts of ill newborns, enriched for African American and rural patients. Our results demonstrate the utility of GS as it provides early in life detection of clinically relevant genetic variation not identified via current standard clinical testing, particularly for newborns exhibiting certain phenotypic features.

## INTRODUCTION

Genome Sequencing (GS) holds tremendous potential value for critically ill newborns with signs suggestive of a genetic disorder. Early diagnosis may be beneficial in the short-term for disease management and treatment but may also shorten the length of the “diagnostic odyssey” that often accompanies rare disease symptoms. Not only is GS capable of detecting a variety of genetic variant types (SNVs, indels, CNVs, aneuploidy), it also allows for assessment of the vast majority of genes in a phenotype-independent manner, a feature that is particularly important when testing very young patients where clinical presentation may not be well-defined until later in life (e.g., intellectual disability). With the discovery of more than 4,000 genes that contribute to Mendelian diseases^1,2^, and with many more yet to be discovered, such comprehensiveness is increasingly valuable.

Although GS is being used to genetically diagnose pediatric patients with rare disease^3-5^, extensive use of GS to diagnose acutely ill newborns is relatively lacking. A few groups have employed GS in a neonatal intensive care unit (NICU) setting^6-8^ and diagnostic rates range from ∼20-50% depending on a variety of factors, including patient selection and testing/analysis methods. While GS holds promise for diagnosing and improving outcomes for ill newborns^9^, many practical questions remain and additional studies on the use of GS as a first-line test are needed; this includes understanding phenotypic features that correlate with diagnostic success rate and which phenotypes may guide future GS usage. Further, it is particularly important to evaluate GS testing in underserved communities such as African Americans, who are sharply under-represented in existing NICU-based translational genomics studies.

SouthSeq is a clinical research study funded as part of the Clinical Sequencing Evidence-Generating Research (CSER) consortium and aims to use GS to detect causal genetic variation in a cohort of NICU newborns with phenotypes suggestive of genetic disease, concurrent with standard of care. A key goal of the study is to evaluate GS as a first-line test to provide an early genetic diagnosis to improve outcomes of affected newborns. SouthSeq targets enrollment of a diverse population of babies representing racial/ethnic minorities as well as those from rural, medically underserved areas. While ongoing, this report describes SouthSeq results from completed analysis of the first 367 probands enrolled across five different clinical sites in the Southeastern US. Our study population of newborns (mean age at enrollment 31 days) is sex-balanced (48% female) and enriched for individuals from diverse and medically underserved populations (74%). All 367 affected babies have received GS and analysis, with 30% receiving a genetic diagnosis and an additional 14% receiving results of uncertain significance. Our results highlight substantial diagnostic utility for GS, as 53% of GS-detected diagnostic variants in SouthSeq were not detected by concurrent standard clinical genetic testing. Moreover, newborns exhibiting abnormal craniofacial, ophthalmologic, auditory and/or skin/hair features were found to be more likely to receive a genetic diagnosis via GS. Finally, we show that although significant technical differences in the interpretation process do exist, diagnostic rates among African American newborns are similar to those observed in European American babies.

## MATERIALS AND METHODS

### Recruitment information

There was no public recruitment for this study. Participant babies were enrolled from the NICU, a high-risk prenatal clinic, or a pediatric unit at one of five SouthSeq clinical sites; University of Alabama at Birmingham/Children’s of Alabama (Birmingham, AL, USA), University of Mississippi Medical Center (Jackson, MS, USA), Woman’s Hospital (Baton Rouge, LA, USA), University of Louisville (Louisville, KY, USA), and Children’s Hospital New Orleans (New Orleans, LA, USA). At least one parent/legal guardian was required to consent for study participation. Antenatal consent was offered to parents when phenotypic features that met enrollment criteria were detected prenatally. Translated consent documents and interpretation services were available for Spanish-speaking participants. SouthSeq employed a custom-built online platform (Genome Gateway) to collect de-identified clinical information for dissemination to the research study team (e.g. consent documentation, demographics, birth history, phenotype, concurrent clinical genetic testing, and opt-in to secondary findings).

### Inclusion/exclusion criteria

For inclusion in SouthSeq, a baby must be inpatient (e.g. neonatal, surgical, cardiac, or pediatric intensive care unit), be in the first 12 months of life (we did enroll one baby who was 379 days old), and exhibit a pattern of congenital anomalies consistent with a genetic disorder and/or present with an unexplained major medical condition (e.g. seizures, metabolic abnormality, etc.). Babies were excluded from the study if they had findings consistent with a known chromosomal aneuploidy (e.g. Trisomy 13, 18, 21 and monosomy X), exhibited anomalies known to result in low diagnostic yield for genetic causes (e.g. isolated gastroschisis, hydronephrosis), or had findings consistent with confirmed teratogenic exposure (e.g. hydantoin, valproate) or congenital infection (e.g. TORCH). Stillborn infants, or babies who died soon after birth, were enrolled if they met inclusion criteria.

### Genome sequencing

Peripheral or cord blood samples collected in EDTA tubes were sent to the HudsonAlpha Clinical Services Laboratory (CSL, a CAP/CLIA-certified genetic testing lab) for DNA extraction (QIAsymphony) and storage. Sequencing libraries were constructed from patient genomic DNA using the CSL’s custom genome library preparation protocol. DNA library fragments were sequenced from both ends (paired) with a read length of 150 base pairs using the Illumina HiSeq X or NovaSeq 6000, with targeted mean coverage depth of 30X and >80% of bases covered at 20X. Sequence reads were aligned to GRCh38 using DRAGEN^10^ or the Sentieon implementation^11^ of BWA-mem. SNVs/indels were called using DRAGEN and GATK^12^ or Strelka^13^.

### GS copy number variant calling

Copy number variants (CNVs) were called from GS bam files using DELLY^14^, ERDS^15^, Manta^16^, and CNVnator^17^. Overlapping calls with at least 90% reciprocity or large calls (>100,000 bp) containing less than 75% segmental duplications were retained if they were observed in eight or fewer unaffected individuals in an in-house database and at less than 1% in population frequency databases (Thousand Genomes^18^, gnomAD^19^). CNVs that survived filtration were subsequently analyzed for potential disease relevance. All rare CNVs found within 5 kb of an established developmental delay/intellectual disability gene, within 5 kb of a MIM^2^ disease-associated gene, or intersecting one or more exons of any gene, were subject to manual curation. CNVs were classified according to ACMG/ClinGen guidelines^20^.

### Annotation, filtering, and variant classification

Identified SNVs and indels were annotated, filtered, and visualized using an in-house software platform. Variants that survived filtration were manually curated and classified as pathogenic, likely pathogenic, or uncertain using ACMG guidelines^21^. Additionally, variants were assigned a case-level designation in the context of clinical presentation that linked variation to confidence in causation, using the terms definitive diagnostic (DD), likely diagnostic (LD), or uncertain. The CSER consortium, spanning a collaborative group of sites performing translational genomic research in a variety of settings, established this case-level classification system and a manuscript describing development and implementation is in preparation. Most newborns harboring pathogenic variation received case-level designation of DD, while those harboring a likely pathogenic variant were classified as LD. However, for a few cases a pathogenic or likely pathogenic variant received case-level designation of uncertain due to insufficient zygosity (only one heterozygous variant was identified in a gene associated with an autosomal recessive or X-linked recessive condition), phenotype mismatch, and/or unknown phase. In contrast, some probands that harbored a variant of uncertain significance received case-level designation of LD based on the specificity of the match between observed and expected phenotypes.

### Variant validation

GS testing was conducted in a CAP/CLIA-certified laboratory, while variant analysis/interpretation was conducted as part of a research protocol. All variants deemed to be returnable to patients/families were clinically tested (via Sanger or array) to confirm variant presence, determine variant inheritance (when parent samples were available), and generate a report with clinical interpretation. Copy number variants were returned in accordance with the research protocol if they were too small to be confirmed via clinical array testing. HGVS nomenclature of all returned variants was verified using VariantValidator^22^.

### Return of Results

Parents or legal guardians of babies enrolled in SouthSeq received GS results via a randomized clinical trial (NCT03842995) comparing standard-of-care return (genetic counselors) to return by trained non-geneticist healthcare providers (NGP; neonatologist, neonatology nurse practitioner). Results of this trial will be published elsewhere. Parents or legal guardians of babies also had the option to receive secondary findings; pathogenic or likely pathogenic variation in an ACMG SFv2.0 gene^23^. GS result reports were placed in the newborn patient’s medical record and follow-up medical care was managed by the appropriate clinical care teams.

## RESULTS

### Demographics

We enrolled 367 babies (365 total families; two families each with two affected babies) with signs suggestive of an underlying genetic disorder (see Methods for study enrollment criteria; Table 1). Patient recruitment occurred at five clinical sites, including the University of Alabama at Birmingham/Children’s of Alabama (UAB, *n*=139, 38%), University of Mississippi Medical Center (UMMC; *n*=118, 32%), Woman’s Hospital in Baton Rouge (BR; *n*=61, 17%), Children’s Hospital in New Orleans (LSU; *n*=31, 8%) and University of Louisville (UL; *n*=18, 5%). UAB and UMMC represent clinical sites where recruitment first began and thus had higher enrollment totals.

**Table 1.** Study demographics

|  | Individuals (n (%)) | Individuals with DD/LD Result (n (%)) |
| --- | --- | --- |
| **Clinical Sites** | **Total (%)** | **Total (%)** |
| University of Alabama at Birmingham and Children's of Alabama (UAB) | 139 (38) | 42 (30) |
| University of Mississippi Medical Center (UMMC) | 118 (32) | 33 (28) |
| Woman's Hospital (BR) | 61 (17) | 20 (33) |
| Children's Hospital in New Orleans (LSU) | 31 (8) | 7 (23) |
| University of Louisville (UL) | 18 (5) | 7 (39) |
| **Sex** | **Total (%)** | **Total (%)** |
| Male | 191 (52) | 54 (28) |
| Female | 176 (48) | 55 (31) |
| **Race/ethnicity** | **Total (%)** | **Total (%)** |
| Black or African American | 126 (34) | 39 (31) |
| White or European American | 187 (51) | 50 (27) |
| More than one category | 34 (9) | 10 (29) |
| Hispanic/Latino(a) only | 13 (4) | 7 (54) |
| Other* | 7 (2) | 3 (43) |
| Racial/Ethnic minority OR medically underserved<sup>#</sup> | 271 (74) | 81 (30) |

| Family Structure | Total (%) |  |
| --- | --- | --- |
| Proband + biological parents | 234 (64) | -- |
| Proband + one biological parent | 104 (28) | -- |
| Proband only | 29 (8) | -- |
| Age | Average (range) |  |
| Age of proband at enrollment (days) | 31 (0-379) | -- |
| Mother age at delivery (years) | 27 (16-49) | -- |
| Birth History | Average (range) |  |
| Gestational age of proband (weeks) | 36 (22-42) | -- |
| Birth weight (g) | 2531.1 (310-5930) | -- |
| Birth length (cm) | 44.97 (20-57) | -- |
| OFC (cm) | 32.3 (18-52.6) | -- |
| Apgar scores (1 min, 5 min) | 5.9 (0-10); 7.5 (0-10) | -- |
| Top HPO terms | Total (%) | Total (%) |
| Abnormality of the face (HP:0000271) | 83 (23%) | 35 (42) |
| Intrauterine growth restriction (HP:0001511) | 80 (22%) | 26 (33) |
| Birth weight less than 10th percentile (HP:0001518) | 64 (18%) | 18 (28) |
| Atrial Septal Defect (HP:0001631) | 59 (16%) | 19 (32) |
| Polyhydramnios (HP:0001561) | 59 (16%) | 20 (35) |

\*Other includes Asian, Middle Eastern, North African or Mediterranean or Unknown, # Based on self-reported income, zip code, race/ethnicity

Participant babies had a mean age of 31 days (range 0-379 days) at time of enrollment and 52% were male. Forty-nine percent of participants (*n*=180) were self-reported non-white and 74% (*n*=271) represent racial/ethnic minorities and/or reside in rural medically underserved areas^24^ (Table 1). With input from NICU providers and medical geneticists, we collected patient clinical information within 13 high-level NICU-relevant phenotypic categories. These categories included 112 pre-selected Human Phenotype Ontology^25^ (HPO) terms (average of seven HPO terms per category), of which 89 were observed in at least one newborn in the study population (Table S1). The five most common HPO terms observed across SouthSeq participants included abnormality of the face (*n*=83, 23%), intrauterine growth restriction (*n*=80, 22%), birth weight less than the 10^th^ percentile (*n*=64, 18%), atrial septal defect (*n*=59, 16%), and polyhydramnios (*n*=59, 16%; Tables 1 and S1).

When available, parental samples were obtained and Sanger tested to assess inheritance of GS-identified potentially clinically relevant variants. Two hundred thirty-four participants were enrolled along with both biological parents, 104 were enrolled with one biological parent, and 29 were enrolled as proband only (Table 1).

### Diagnostic yield

We conducted GS to identify genetic variation associated with rare congenital phenotypes. Sequencing reads were generated using Illumina sequencers with a mean genome-wide coverage of 35X across all probands. SNVs, indels, and CNVs were annotated and filtered using a variety of technical and biological features and subjected to ACMG-guided classification^21^. Variants deemed returnable after manual curation were sent to an independent CAP/CLIA laboratory for clinical Sanger confirmation and reporting. In addition to the clinical genetics report, participant families were provided with a letter, written by a genetic counselor, explaining variant interpretation in the context of phenotype information. Results were disclosed to families by either a genetic counselor or a non-genetics provider trained to return GS results by the SouthSeq study team (see Methods).

Of the 367 sequenced probands, 160 (44% of our cohort) received a result associated with the primary indication for testing. Results designated as definitive diagnostic (DD) or likely diagnostic (LD) were identified in 109 babies (30%), while an additional 51 participants (14%) received an uncertain case-level result (Figure 1A; Table S2). Returnable variation was identified across 107 genes (64 harboring DD/LD findings, 43 harboring an uncertain finding). Eight genes were found to harbor unique DD/LD variants in two or more unrelated probands (Tables S2 and S3), with six babies genetically diagnosed with CHARGE syndrome (*CHD7*, MIM:214800) and six with Noonan syndrome (*PTPN11*, MIM: 163950). Turnaround time from enrollment to report generation averaged 73 days, which included ∼30 days for Sanger confirmation in an independent CAP/CLIA laboratory.

**Figure 1.**
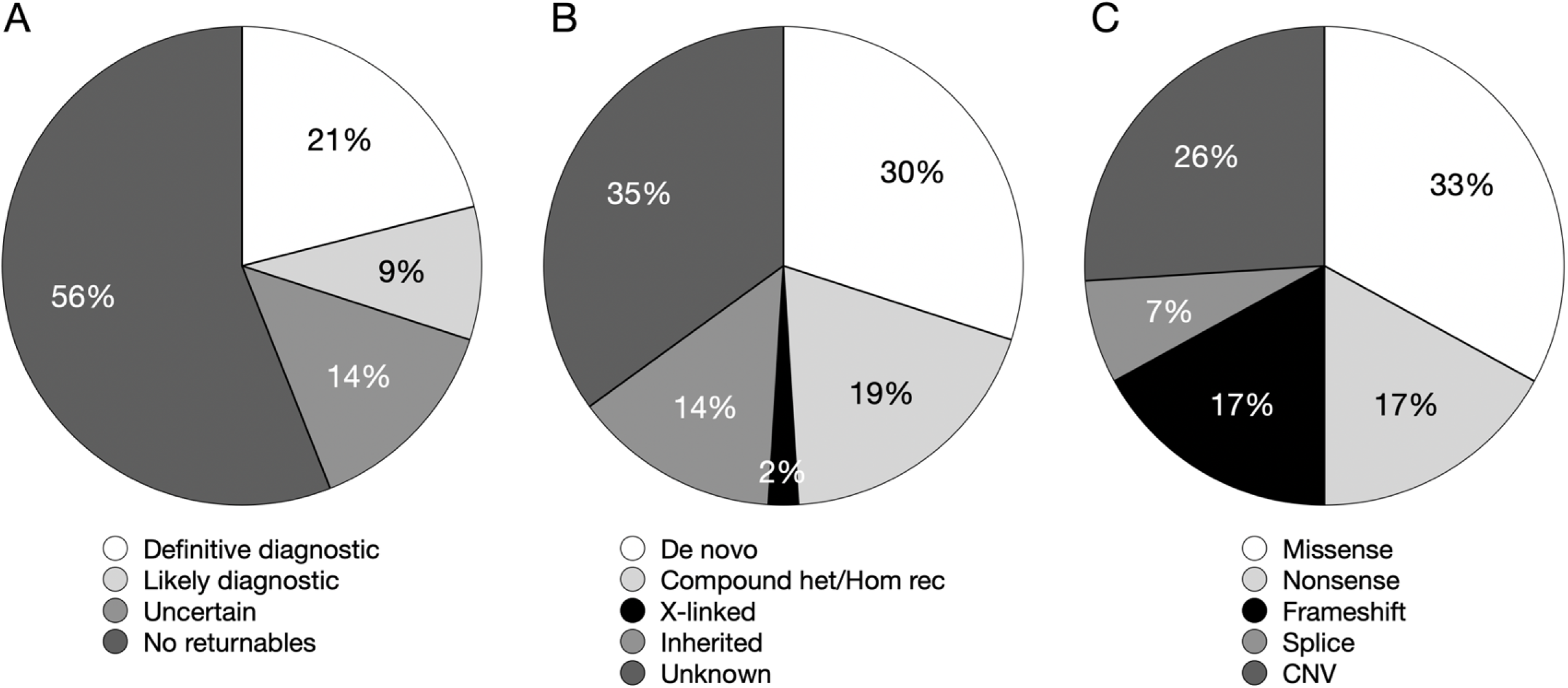
Utility of genome sequencing as a first-line genetic test for newborns (n=367 babies). (A) Diagnostic yield of SouthSeq study population. Thirty percent (n=109) of study participants received a definitive diagnostic (DD) or likely diagnostic (LD) finding; 14% (n=51) received an uncertain finding, and no genetic findings of interest were identified in the remaining 56% (n=207). (B) Percentage of findings that fall within each mode of inheritance category, including *de novo*, compound heterozygous or homozygous, X-linked, inherited, or unknown. Unknown represents heterozygous variants (SNV or CNV) where one or both parents were unavailable for testing, or inheritance could not be determined (e.g. non-paternity). (C) Types of variation represented by DD/LD findings, including missense, nonsense, frameshift, splice, and CNV; one case of uniparental disomy (UPD) not shown.

Additionally, 89% percent of families consented to receive secondary results in an ACMG SFv2.0 gene^23^ with seven pathogenic/likely pathogenic (P/LP) variants detected across six newborns (1.6%). One baby harbored two P/LP findings in two different genes, *MSH6* and *ACTC1* (Table S4).

In 30% of DD/LD cases the returnable variant occurred *de novo* in a dominant disease gene, while 19% of DD/LD newborns inherited compound heterozygous or homozygous variation in a recessive disease gene. Fourteen percent of DD/LD newborns inherited a heterozygous variant in a dominant disease gene from a parent, some of which were affected (Figure 1B). An additional 2% of DD/LD cases were maternally inherited X-linked recessive findings in males. Finally, 35% of DD/LD cases were of unknown inheritance due to biological parent samples being unavailable (Figure 1B). DD/LD findings represent a variety of variant types; 33% of identified DD/LD variants result in missense, 17% in nonsense, and 17% in frameshift. Further, 7% were predicted to disrupt splicing (at or near a canonical splice site), and 26% represent a copy number variant (CNV; Figure 1C). There was also one DD case resulting from uniparental disomy.

### Factors that drive diagnostic yield

We compared available clinical, demographic, and phenotypic variables to identify potential correlations with diagnostic yield, including site of enrollment, sex, race/ethnicity, access to care, and phenotype (Tables 1 and 2).

**Table 2.** Diagnostic yield and phenotype. GS testing may yield increased diagnoses for patients with certain phenotypic features. Odds ratios and p-values calculated using Fisher’s exact test (* denotes statistical significance).

| Phenotype categories | # Probands | # Probands with DD/LD (%) | P-value (odds ratio) | # Probands with Uncertain (%) | P-value (odds ratio) |
| --- | --- | --- | --- | --- | --- |
| **Study-wide totals** | 367 | 109 (30) | --- | 51 (14) | --- |
| **Prenatal** | 148 (40) | 50 (34) | 0.16 (1.40) | 21 (14) | 0.88 (1.04) |
| **Craniofacial/Ophthalmologic/Auditory** | 144 (39) | 60 (42) | <0.0001 (2.54)* | 23 (16) | 0.36 (1.32) |
| **Cardiac/Congenital heart malformations** | 142 (39) | 44 (31) | 0.73 (1.11) | 18 (13) | 0.64 (0.84) |
| **Growth** | 139 (38) | 43 (31) | 0.72 (1.09) | 15 (11) | 0.21 (0.65) |
| **Skeletal/Limb abnormalities** | 99 (27) | 34 (34) | 0.25 (1.35) | 18 (18) | 0.17 (1.58) |
| **Neurological/Muscular** | 91 (25) | 25 (28) | 0.69 (0.87) | 22 (24) | 0.003 (2.72)* |
| **Genitourinary abnormalities** | 86 (23) | 31 (36) | 0.17 (1.47) | 14 (16) | 0.48 (1.28) |
| **Brain malformations/abnormal imaging** | 66 (18) | 21 (32) | 0.66 (1.13) | 10 (15) | 0.70 (1.13) |
| **Gastrointestinal** | 50 (14) | 9 (18) | 0.06 (0.48) | 8 (16) | 0.66 (1.21) |
| **Hematologic/Immunologic** | 36 (10) | 7 (19) | 0.25 (0.56) | 2 (6) | 0.20 (0.34) |
| **Metabolic** | 30 (8) | 9 (30) | 0.99 (1.05) | 4 (13) | 0.99 (0.95) |
| **Skin/Hair** | 27 (7) | 13 (48) | 0.03 (2.43)* | 4 (15) | 0.78 (1.08) |
| **Endocrine** | 7 (2) | 0 (0) | 0.11 (0.00) | 0 (0) | 0.60 (0.00) |

We observed no difference in diagnostic yield between the two primary clinical sites that account for most study enrollments (30% at UAB vs. 28% at UMMC), and the rates at the other three nurseries are similar albeit more variable due to smaller enrollment totals (33%, 23%, and 39% at Woman’s Hospital, Children’s Hospital in New Orleans, and University of Louisville, respectively), suggesting roughly similar overall diagnostic yields across these NICU locations (Table 1).

We observed no difference in diagnostic yield between self-reported African American (AA) and European American (EA) babies (31% vs 27%, respectively). However, analysis of AA genomes required more manual curation and Sanger testing than EA genomes. After primary variant filtration, ∼1.4X more variants were retained and required manual curation for an AA genome (average of 300 variants/proband, n=111 genomes) compared with an EA genome (average of 221 variants per proband, n=160 genomes). Additionally, we Sanger tested parent samples to determine inheritance or phase for candidate variants prior to final interpretation given that some ACMG evidence codes require such information. For EA babies that were enrolled with both biological parents, we averaged 0.79 Sanger tests/proband (116 Sanger tests across 146 EA probands) compared with 0.96 Sanger tests per proband for AA babies (54 Sanger tests across 56 AA probands; p=0.033, two-proportion z-test).

We also examined diagnostic yield in the context of phenotype. As previously mentioned, SouthSeq-associated NICU providers and medical geneticists provided patient phenotype information within 13 high-level categories containing 112 NICU-relevant HPO terms (Tables 2 and S1). Because any given baby may exhibit more than one phenotype (HPO term) in each category, we conducted calculations based on total unique individuals in each. Phenotypic categories that were attributed to the largest number of participants included prenatal (e.g., IUGR, amniotic fluid levels, cystic hygroma; n=148, 40%), craniofacial/ophthalmologic/auditory (n=144, 39%), cardiac/congenital heart malformations (n=142, 39%), growth (e.g., birth weight/length, head circumference, failure to thrive; n=139, n=38%), and skeletal/limb abnormalities (n=99, 27%; Table 2).

We found substantial variation in DD/LD and uncertain yield across phenotypic categories. For example, DD/LD rates ranged from 18% among babies with gastrointestinal findings to 48% for those with skin/hair findings. We also compared DD/LD or uncertain rates between babies who do and do not exhibit phenotypic features across each category. We found that babies with craniofacial/ophthalmologic/auditory abnormalities had the largest and most significant enrichment for DD/LD findings (OR=2.54, p<0.0001), followed by skin/hair abnormalities (OR=2.43, p=0.03). Our data also suggest that babies with neurological and/or muscular findings (OR=2.72, p=0.003) are more likely to receive an uncertain result (Table 2). We found no phenotypic category that is significantly depleted of DD/LD findings, although we observed a substantially lower diagnostic yield for babies who presented with abnormal gastrointestinal features (OR=0.48, p=0.06).

### Utility of genome sequencing

The SouthSeq study design explicitly stated that standard clinical care and testing should be conducted regardless of SouthSeq participation. Reflecting this, the vast majority (86% percent, n=314 of 367) of SouthSeq babies received at least one clinical genetic test in parallel with SouthSeq GS, with an average of 1.7 (range 0-5) tests per baby. Most babies received clinical CNV testing (75%, n=234), while many received postnatal karyotype/FISH (39%, n=124), prenatal screening or testing (42%, n=132; e.g., quad screen, amniocentesis, maternal serum screen, noninvasive prenatal screening (NIPS)) or postnatal single-gene/panel testing (26%, n=81). A small fraction received clinical exome testing (3%, n=11; Table S5).

Thirty-nine percent (43 of 109) of GS-detected DD/LD findings were also identified by clinical genetic testing (Tables 3, Table S5). Among these, 58% (25 of 43) were identified by CNV testing, 37% (16 of 43) by gene panel, and 5% (2 of 43) by clinical exome. Nearly 16% (8 of 51) of GS-detected uncertain results were also identified by clinical genetic testing (Tables 3, Table S5), mostly via CNV testing (88%, n=7). For 20/51 babies where GS and clinical genetic testing identified the same genetic variant (including uncertain cases), SouthSeq was able to provide families with inheritance information (including nine *de novo* and 11 inherited variants) not generated by standard testing.

**Table 3.**
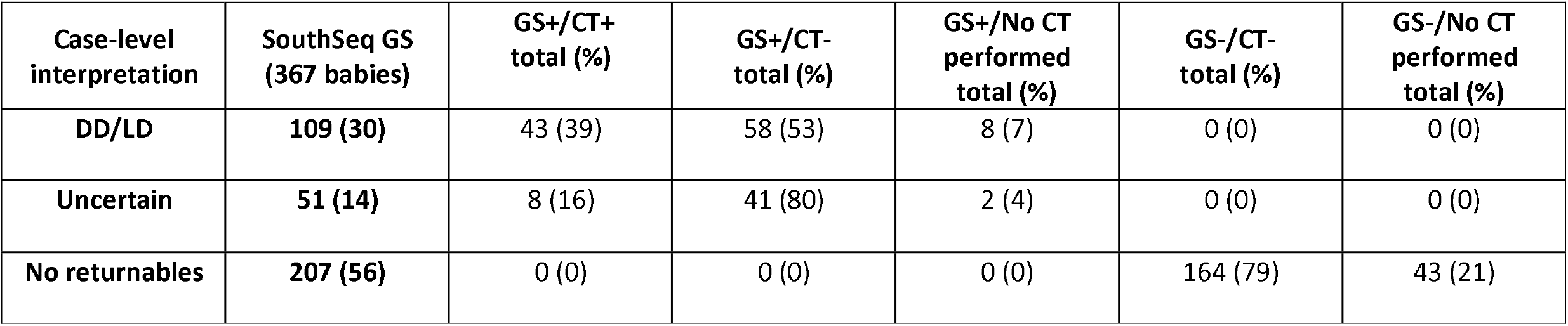
Overlap of findings between GS findings and clinical genetic testing (GS, genome sequencing; CT, clinical genetic testing; CT-does not include babies that did not receive any clinical genetic testing).

Most GS-detected DD/LD events were not detected via standard testing (Tables 3 and S5). Reasons that clinical genetic testing missed diagnoses include 1) GS detected a DD/LD SNV or indel but no single gene/panel test was ordered (43 of 58 cases); 2) GS detected a DD/LD SNV or indel in a gene not on a clinically ordered single gene/panel test (13 of 58 cases); or 3) GS detected a DD/LD CNV when clinical CNV testing was not ordered (2 of 58 cases). Case studies that highlight DD/LD results missed by clinical genetic testing, and that demonstrate the benefit of GS testing in newborn patients, are provided in Table 4.

**Table 4.** Cases studies highlighting the utility of genome sequencing as a first-line genetic test.

| Participant | SS-47 | SS-231 | SS-51 |
| --- | --- | --- | --- |
| Clinical phenotype | Open lip schizencephaly, IUGR, congenital microcephaly, external ear malformation, central hypotonia, possible dysgenesis of corpus callosum, malformed large right ventricle, gliosis, calcification in right brain hemisphere and large right ventricle, high arched hard palate, poor head control, absent Moro reflex, upgoing toe signs, ankle clonus; TORCH suspected | Polyhydramnios, fetal bradycardia, open and flat anterior fontanelle, upturned nasal tip, tented upper lip, high arched palate, posterior hair whorl, central hypotonia, downslanted palpebral fissures, muscular hypotonia, low-set ears, hand clenching, poor reflexes, required intubation/failed extubation | IUGR, low set ears, undescended testis, jejunal atresia, mild cardiomegaly, rocker bottom feet, bilateral 5th finger clinodactyly, moderate petechiae on back, buttocks, and flank, café au lait spots in lumbar/sacral area, abnormal EEG suggestive of seizure activity |
| Clinical testing completed | TORCH screening, CMV testing, CNV testing - all non-diagnostic | Karyotype, CNV testing, SMA testing, methylation testing, gene panel testing (x2), repeat expansion testing, mitochondrial genome panel, brain MRI, muscle biopsy, biochemical studies - all non-diagnostic | Karyotype, CNV testing, seizure panel – all non-diagnostic |
| Findings of SouthSeq GS | NM_001845.5(*COL4A1*):c.3556G>A (p.G1186S) | NM_003632.2(*CNTNAP1*):c.1801_2292+647del;<br>NM_003632.2(*CNTNAP1*):c.2901_2902del (p.C968Ffs*11) | NM_001020658.1(*PUM1*):c.3439C>T (p.R1147W) |
| Associated disease (MIM#) | Brain small vessel disease with or without ocular anomalies (175780) | Lethal congenital contracture syndrome 7 (618186); Hypomyelinating neuropathy, congenital 3 (616286) | Spinocerebellar ataxia 47 (617931) |
| Variant inheritance | *de novo* | unknown; paternal (variants in trans) | *de novo* |
| Variant-level classification | Likely pathogenic (PS2, PM2, PP3) | Pathogenic (PVS1, PM2, PM3);<br>Pathogenic (PVS1, PM2, PM3, PP1S) | Pathogenic (PS2, PS3, PS4M, PM2, PP3) |
| Case-level classification | Likely diagnostic | Diagnostic | Diagnostic |

Of the 51 GS-identified uncertain findings, 41 (80%) were not detected by standard testing, likely owing at least partially to the inherent uncertainty related to these findings (e.g., questions about phenotype match, uncertainty of variant impact, etc.). Nine of the GS uncertain cases included a likely deleterious variant in a candidate gene not currently associated with disease, but which may become a disease gene in the future (e.g., via GeneMatcher submission^26^). An additional nine uncertain GS results arose from identification of a single P/LP variant in a gene associated with an autosomal recessive condition for which there was significant phenotypic overlap. While we did return such variation in SouthSeq, classified as case-level uncertain due to insufficient zygosity, none of these variants were returned via standard clinical testing.

Thirty-two VUSs were detected via clinical genetic testing and, although detected via GS, were not returned by the study (Table S5). Most of these participants were GS negative (n=18), while nine had GS-identified DD/LD findings, and five had GS-identified uncertain findings that did not match the variants returned via clinical testing. Reasons for discrepancies in return between the clinical lab and SouthSeq include: 1) variant in a highly penetrant disease gene inherited from an unaffected parent; 2) phenotype mismatch; 3) observed too frequently in population frequency databases; 4) VUS in a gene associated with an autosomal recessive condition without a match to observed phenotypes.

## DISCUSSION

In SouthSeq, we used GS, concurrently with standard of care, in a diverse cohort of infants from underserved and rural populations who presented with phenotypic features suggestive of a genetic disorder. Sanger testing was completed in parent samples (when available) to determine variant inheritance and aid interpretation. 44% of newborn SouthSeq participants were found to harbor a genetic variant associated with indication for testing; 30% received a DD/LD result, and an additional 14% received a result of uncertain significance. This observed diagnostic rate is in line with previous reports from other groups^6-8^. Returnable findings include different variant types (SNVs, indels, CNVs) affecting 107 unique genes, highlighting the comprehensiveness of GS. Finally, we identified seven P/LP variants across six babies (one baby with two findings, 1.6%) in an ACMG SFv2.0 gene.

We observed no significant difference in diagnostic rate between the two largest racial/ethnic categories in SouthSeq (31% diagnostic rate for AA babies, 27% for EA babies). AA babies did, however, require more manual curation of variants that appeared to be rare and damaging, and also required more Sanger testing in parent samples to determine variant inheritance (0.79 Sanger tests/EA trio babies, 0.96 tests/AA trio babies). These results are consistent with previous reports of ancestry-associated differences in clinical genetic testing procedures and are likely to result, at least in part, from under-representation of African alleles in population frequency databases^27,28^. Increased population frequency data for minority populations would likely reduce this discrepancy.

Because the resources necessary to conduct GS are often limited, identification of those patients most likely to benefit from GS testing could maximize GS utility. As such, we sought to identify patient attributes that correlate with the likelihood of receiving a genetic diagnosis (Table 1). We grouped newborn participants into 13 high-level phenotype categories based on 112 pre-defined HPO terms and compared rates of different finding types (DD/LD and uncertain) between babies that exhibit features falling within a specific category and those that do not (Table 2). We found that newborns that exhibited abnormal craniofacial, ophthalmologic, auditory, skin, or hair findings were ∼2.5X more likely (p<0.0001) to receive a DD/LD result compared with babies who did not exhibit these phenotypic attributes. Conversely, although not statistically significant, babies with gastrointestinal phenotypes received a DD/LD result at ∼50% the rate of babies that did not exhibit gastrointestinal abnormalities. Finally, individuals with neurological/muscular-associated features were 2.7X (p=0.003) more likely to receive a variant of uncertain significance. This observation may reflect the complexities of variant interpretation, particularly for neurological features that may not be assessable in infancy^29^. Although others have conducted similar analyses^30,31^, future larger studies may help to further delineate clinical features most predictive of GS diagnostic potential.

In SouthSeq, enrolled babies received GS concurrent with standard of care, including clinical genetic testing. Because of this, we were able to compare the results from standard clinical genetic testing with GS. We found that most GS DD/LD results, affecting 18 % of all enrolled babies, were not detected via standard of care; conversely, no DD/LD findings from standard testing were missed by GS, although it should be noted that positive clinical testing results were made available to the SouthSeq study team for a few cases at time of GS analysis. As would be suspected, most of the DD/LD variation missed by standard testing resulted from the fact that the clinical tests ordered were not capable of detecting the missed relevant variants. In contrast to single-gene or gene-panel tests, for example, GS testing is independent of hypotheses about which specific disease genes may be involved. Further, early use of GS can reduce the number of tests required to obtain a diagnosis; for example, on average almost two in-parallel non-GS genetic tests were ordered per SouthSeq baby despite leading to an overall yield less than half that of GS. As such, GS has considerable potential to prevent the multiplicity of testing associated with the “diagnostic odyssey”^32,33^ that many rare-disease patients experience.

In conclusion, we have conducted GS testing for a cohort of infants affected with suspected congenital anomalies and enriched for individuals from medically underserved and historically underrepresented groups in genomics research. We show that newborns with certain phenotypic features may benefit more from GS testing and highlight the comprehensiveness of GS and its benefit beyond standard clinical genetic testing. Our data strongly support using GS as a first-line genetic test for seriously ill newborns.

## Data Availability

Genome sequencing data for all participants is in the process of being shared through the NHGRI Analysis Visualization and Informatics Lab-space (AnVIL). All reported variants have been submitted to ClinVar.

## Data availability

The sequencing data generated by this work are available through AnVIL/dbGaP^34^ (phs002307.v1.p1) and interpreted variants have been placed into ClinVar^35^ under the study name ‘CSER-SouthSeq’.

## Acknowledgements

We are grateful to the patients and their families who contributed to this study. We thank the HudsonAlpha Software Development and Informatics team and the Clinical Services Laboratory who contributed to data acquisition and analysis. The SouthSeq project (U01HG007301) is supported by the Clinical Sequencing Evidence-Generating Research (CSER) consortium which is funded by the National Human Genome Research Institute (NHGRI) with co-funding from the National Institute on Minority Health and Health Disparities (NIMHD) and the National Cancer Institute (NCI). More information about CSER can be found at https://cser-consortium.org/.

## Author Information

Conceptualization: G.M.C., B.R.K., G.S.B.; Data curation; K.M.B., M.L.T., C.R.F., S.M.H., D.R.L., M.D.A., J.M.J.L, D.E.G.; Formal analysis: K.M.B., M.L.T., C.R.F., S.M.H., D.R.L., M.D.A., J.M.J.L., D.E.G.; K.M.E., M.E.C., V.G., W.V.K.; Funding acquisition: G.M.C., B.R.K., G.S.B.; Investigation: K.M.B., M.L.T., C.R.F., S.M.H., D.R.L., M.D.A., J.M.J.L., K.M.E., M.E.C., V.G., W.V.K., D.E.G., S.A.F., H.M., A.C., A.L., K.E.J., L.G.M., H.M.J., M.J., L.A.M., S.L.D., C.T., H.W., K.L., M.B.N.; Methodology: K.M.E., M.E.C., V.G.; Project administration: K.M.B., M.L.T., C.R.F., K.M.E., M.E.C., V.G., W.V.K., H.M., A.C., A.L., K.E.J., L.G.M., H.M.J., M.J., L.A.M., S.L.D., C.T., H.W., K.L.; Software: J.M.J.L, D.E.G.; Supervision: K.M.B., C.R.F., K.M.E., J.P.E, A.C.E.H., J.K., W.C., K.B.B., B.M.K., R.S., D.S., S.B.S., S.J.K., G.S.B., B.R.K., G.M.C.; Validation: K.M.E., M.E.C., V.G., W.V.K.; Writing-original draft: K.M.B., M.L.T., C.R.F., J.M.J.L., D.R.L., K.M.E.; Writing-review & editing: K.M.B., M.L.T., C.R.F., S.M.H., D.R.L., J.M.J.L, K.M.E., M.E.C., S.A.F., M.J., J.P.E, A.C.E.H., K.B.B., B.M.K., R.S., S.B.S., S.J.K., G.S.B., B.R.K., G.M.C

## Ethics Declaration

All authors declare no competing financial interests in relation to the work described. The review board at the University of Alabama at Birmingham (IRB-300000328) approved and monitored the study. All study participants (parent or legal guardian) were required to give written consent to participate in the study. All individual-level data was de-identified to the research team. The authors received and archived written patient consent to publish individual data.

