## Supplemental Table 3 for "Genome sequencing as a first-line diagnostic test for hospitalized newborns"

Table S3. Recurrent diagnostic gene findings across 367 affected newborns

| Number of cases (*n*) | Gene | OMIM Disease (Inheritance) |
| --- | --- | --- |
| 6 | *CHD7* | CHARGE syndrome (AD) |
| 6 | *PTPN11* | Noonan syndrome 1 (AD) |
| 3 | *KMT2D* | Kabuki syndrome (AD) |
| 3 | *MYRF* | Cardiac-urogenital syndrome (AD) |
| 2 | *COL2A1* | COL2A1-related disorders (AD) |
| 2 | *DNAH5* | Ciliary dyskinesia, primary, 3, with or without situs inversus (AR) |
| 2 | *TFAP2A* | Branchiooculofacial syndrome (AD) |
| 2 | *TP63* | Ectrodactyly, ectodermal dysplasia, and cleft lip/palate syndrome 3 (AD) |
